# Segmental MRI Pituitary and Hypothalamus Volumes post Fontan: An analysis of the Australian and New Zealand Fontan Registry

**DOI:** 10.1101/2024.07.05.24309972

**Authors:** Waverley Gee, Joseph Yuan-Mou Yang, Tom Gentles, Sonja Bastin, Ajay J Iyengar, Jian Chen, Dug Yeo Han, Rachael Cordina, Charlotte Verrall, Craig Jefferies, The Australian and New Zealand Fontan Registry

## Abstract

**Objective:** Short stature, central hypothyroidism and infertility are common in those with a Fontan circulation. Given that the Fontan circulation often results in hepatic portal venous congestion, we hypothesize that the hypothalamic-pituitary portal circulation is also affected, contributing to subsequent hypothalamic-pituitary axis dysfunction.

**Methods:** MRI data from the Australian and New Zealand Fontan Registry (86 cases) was compared to 86 age- and sex- matched normal published controls. Total pituitary volumes (both anterior and posterior glands) were measured using a manual tracing segmentation method, and hypothalamic (and subunit) volumes using an automated segmentation tool. Measured gland volume was normalized to total brain volumes. A generalized linear model was used for statistical analysis.

**Results:** Normalized total pituitary volumes (nTPV) were increased in Fontan patients compared to controls (*p* <0.0001), due to an increase in anterior pituitary volumes (nAPV) (*p* <0.0001), with no difference in normalized posterior pituitary volumes (*p* = 0.7). Furthermore, normalized anterior and tubular hypothalamic subunit groups) were increased in Fontan patients compared to the controls (*p* <0.01 and *p* <0.0001, respectively).

The time between Fontan and MRI was positively related to nTPV, nAPV and bilateral hypothalamic volumes. nTPV increased with age, and the increase in nAPV was greater in Fontan patients.

**Conclusions:** Segmental MRI Pituitary and Hypothalamus volumes post Fontan are increased and are related to the time since Fontan procedure. These findings are consistent with venous congestion of the anterior hypothalamic-pituitary portal venous system and may explain the high frequency of endocrine dysfunction in this patient group.

## Introduction

The Fontan procedure is the destination treatment for children born with a functional single ventricle. It results in diversion of systemic venous blood directly to the pulmonary arteries such that there is no sub-pulmonary ventricle. Consequently, the central venous pressure is elevated, and cardiac output reduced. Hepatic portal venous congestion is common, as is progressive liver disease. ^1^

The special characteristic of the hypothalamus is that it connects to the pituitary gland by two different mechanisms: by neuroendocrine neurons projecting directly to the posterior lobe of the pituitary, or by portal vessels to the anterior lobe of the pituitary.^2^ Thus the anterior pituitary is a portal venous system and at potential risk of congestion as per the hepatic portal system. Hypothalamic-pituitary-axis (HPA) dysfunction may be more common than realized in post Fontan circulation given the high frequency of short stature, central hypothyroidism and hypogonadic hypogonadism reported in this unique patient population. ^3–6^ One previous study has demonstrated significantly larger pituitary volumes in children post Fontan compared to controls, however the sample size was small, and methodology did not allow segmental volumetrics. To date these findings have not yet been replicated. ^77^There have also been no studies to date investigating the hypothalamus volumetrics in Fontan patients. Given the vital role this structure plays in the HPA axis and its differential (neuroendocrine or portal venous) anatomical link to the anterior and posterior pituitary gland, assessing its volumetrics may provide more knowledge on the effect Fontan circulation has on the HPA.

Our study aimed to compare both the pituitary and hypothalamus volumes from the Australian and New Zealand (ANZ) Fontan Registry with published normative controls. The ANZ Fontan Registry is a large population-based Fontan registry and provides an opportunity to replicate aspects of the previous study in a larger population. We hypothesize that anterior (and therefore the total) pituitary volumes are increased in Fontan patients compared to normative controls. We were able to utilize a segmentation protocol to MR images, thus increasing the accuracy of measurement and allowing quantification of anterior and posterior gland volumes separately. We also hypothesize that hypothalamus volumes related to the anterior pituitary are increased in patients with a Fontan circulation due to portal venous congestion of the hypothalamic-pituitary axis.

## Methods

### Participants

Patients with a Fontan circulation were recruited as part of the Australia and New Zealand Fontan Registry ‘Functional Outcomes after Fontan Surgery’ Study between April 2015 and November 2018. ^8^ Recruitment occurred across four sites: The Children’s Hospital at Westmead, Sydney; Royal Prince Alfred Hospital, Sydney; The Royal Children’s Hospital, Melbourne; Starship Children’s Hospital, Auckland. The study protocol was approved by the appropriate institutional review board for each site, and written informed consent was obtained from all participants.

Inclusion criteria encompassed individuals with a Fontan circulation who were between 13 and 49 years of age, ≥5 years post-Fontan completion and who were enrolled in the Australian and New Zealand Fontan Registry.

Exclusion criteria included those that had a contraindication to undergo MRI (therefore had no MRI data available) and/or an existing diagnosis of severe intellectual disability or a clinically diagnosed genetic syndrome associated with cognitive impairment identified from their medical record.

### Controls

MRI brain data was acquired on a 1:1 basis matching each Fontan participant for age and sex. This data was obtained from three public-access brain MRI databases: Autism Brain Imaging Data Exchange (ABIDE) I and II (https://fcon_1000.projects.nitrc.org/indi/abide/) and Pediatric Imaging, Neurocognition, and Genetics (PING) data repository (http://pingstudy.ucsd.edu). ^9–11^ ABIDE I and II subjects were selected from the ‘typical controls’ pool: people without neurological or neurocognitive disorder or congenital heart disease. PING participants were a ‘normal’ cohort without neurological or neurocognitive disorders.

### Data Collection

#### Brain MRI

Brain MRI data was acquired using Siemens 3Tesla MRI scanners at each site.^8^ The sequences utilized for segmentation included 3D T1- and T2-weighted imaging. Further details can be found in the supplementary material.

One hundred Fontan cases in the registry had MRI brain imaging. Eighty-six of these had sufficient scan quality (fourteen failed image processing). Demographics and other characteristics of these subjects and of matched controls are detailed in Table 1.

**Table 1.** Summary of the five hypothalamic subunits and the included nuclei.

| Subunits | Nuclei included |
| --- | --- |
| Anterior-superior (a-sHyp) | Preoptic area; paraventricular nucleus |
| Anterior-inferior (a-iHyp) | Suprachiasmatic nucleus; supraoptic nucleus |
| Superior tubular (supTub) | Dorsomedial nucleus; paraventricular nucleus; lateral hypothalamus |
| Inferior tubular (infTub) | Arcuate (or infundibular) nucleus; ventromedial nucleus; supraoptic nucleus; lateral tubular nucleus; tuberomammillary nucleus |
| Posterior (posHyp) | Mamillary body (including medial and lateral mamillary nuclei); lateral hypothalamus; tuberomammillary nucleus and posterior nucleus |

(Adopted and modified from Billot 2020, CC by 4.0 DEED copyright permission.)

3D T1-weighted data was pre-processed using the standard ‘recon-all’ pipeline in the FreeSurfer image analysis suite (version 6.0; https://surfer.nmr.mgh.harvard.edu/). ^12,13^ This pipeline performs intensity non-uniformity correction and normalization, skull stripping, resampled all T1-weighted images to 1.0 mm isotropic resolution. Total brain volume (TBV) was calculated for each participant using the ‘*BrainSe*g’ volume output from the ‘*recon-all’* pipeline. The TBV was used to normalize the pituitary and hypothalamus subgroup volume.

#### Pituitary gland segmentation protocol

The pituitary gland was segmented by manually tracing the structure of 3D T1-weighted images using the MRtrix3 software. This was performed by Dr JYMY, a neurosurgery research fellow with expert neuroanatomic knowledge, and over 10 years of advanced neuroimaging research experience. We adopted the segmentation protocol described by Farrow et al., 2020.^14^ This protocol was chosen based on the simplicity of its anatomical descriptions, and excellent intra- and inter-rater reliability demonstrated by all previous studies adopted this protocol for pituitary volumetric.^15–20^ Please refer to the supplementary material for details of the protocol from Farrow.

The total pituitary gland volume (TPV) is the sum of the anterior pituitary gland (APV), and posterior pituitary gland (PPV) volumes. All volumes were calculated by summing the voxel volumes within each respective traced region.

For New Zealand site participants (n = 21), the sellar region anatomy was not able to be clearly visualized based on the type of 3D T1-weighted data (MP2RAGE) used. 3D T2- weighted data was used instead. It was pre-processed the same way as the T1-weighted data (intensity non-uniformity correction and normalization, resampled to 1.00 mm isotropic resolution). The T2-weighted data did not have the image contrast required to differentiate anterior and posterior pituitary glands; thus, only total pituitary volume was calculated for these cases.

#### Hypothalamus and subunit segmentation protocol

Manual delineation of the hypothalamus and its nuclei suffers from inaccuracy and reproducibility issues due to lack of T1-weighted image contrast within the hypothalamus.

We adopted an automated tool based on a deep convolutional neural network (CNN; https://github.com/BBillot/hypothalamus_seg) for segmentation of the hypothalamus and its subunits (anterior-superior, anterior-inferior, superior tubular, inferior tubular and posterior) from T1-weighted MRI scans. ^21^ The training data for this CNN network was based on previously described manual delineation of hypothalamus and its five hypothalamic subunits in healthy adults (Figure 2), using anatomical landmarks visible on standard 3D structural T1-weighed images. ^22,23^ It has been demonstrated that both the whole hypothalamus and the five hypothalamic subunits can be reliably segmented at standard 1.0 mm resolution using this method. ^22,23^ This schema of morphometric parcellation of the human hypothalamus is similar to the classical approach of subdivision of this structure; parcellating it into three general groups (anterior, tubular and posterior).^21^ Figure 2 and Table 1 demonstrate example parcellation images and detailed anatomical descriptions.

**Figure 1.**
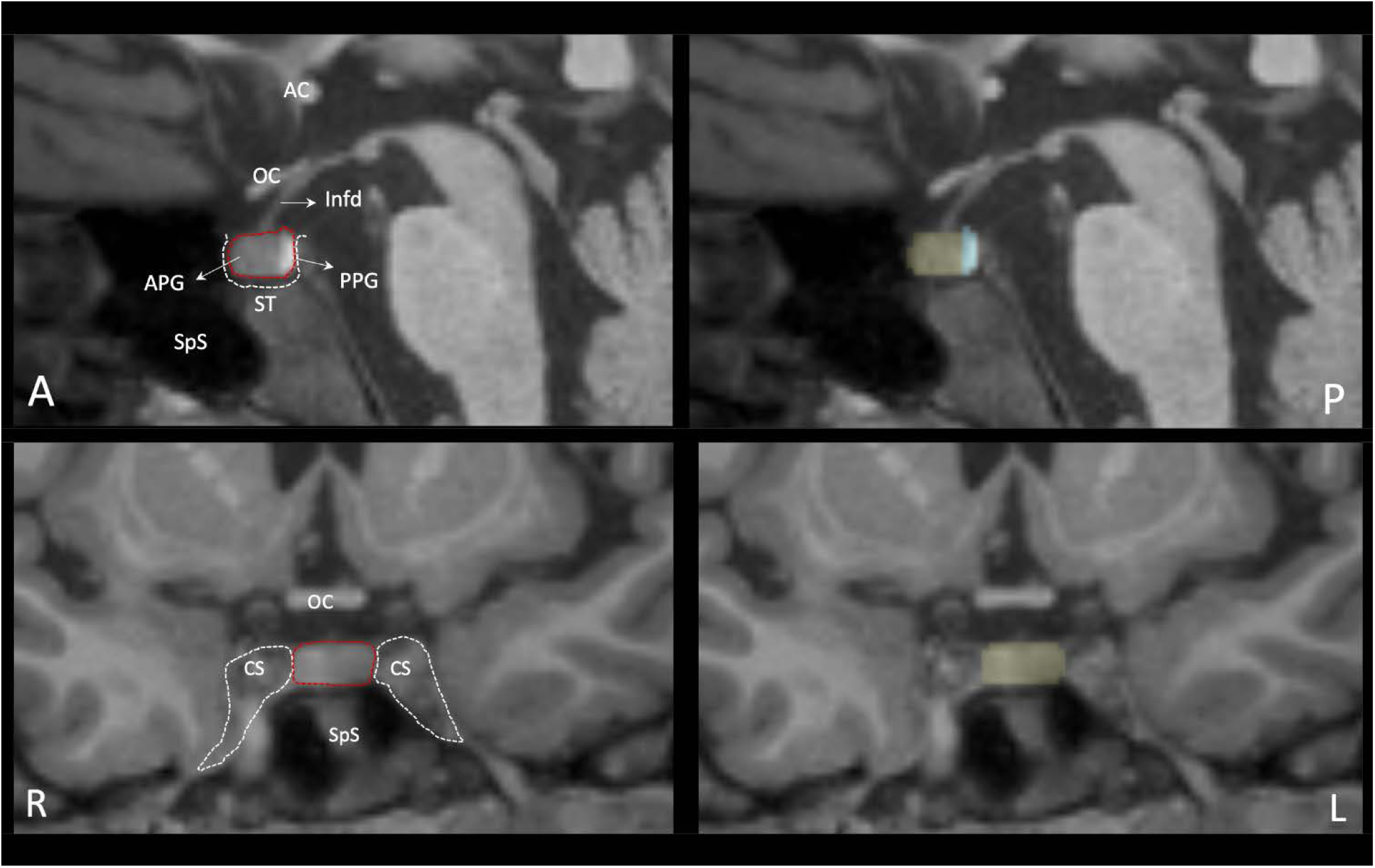
Pituitary gland anatomy and segmentation protocol on T1-weighted MRI. The pituitary gland is identified in the mid-sagittal plane, located within the *sella turcica* (ST) of the sphenoid bone. It is superiorly bounded by the *diaphragm sellae*, laterally bounded by the *cavernous sinuses* (CS) and inferiorly bounded by *the sphenoid sinus* (SpS). It has an *anterior gland* (APG), a p*osterior gland* (PPG), which is a direct caudal extension of the hypothalamus via the *infundibulum* (Infd.). The pituitary stalk is formed by the infundibulum and *pars tuberalis* of the APG. The T1W hyperintensity displayed by the PPG helps separate tracing of the APG (in yellow) and PPG (in cyan). Other abbreviations: AC = anterior commissure; OC = optic chiasm; A = anterior; P = posterior; R = right; L = left.

**Figure 2:**
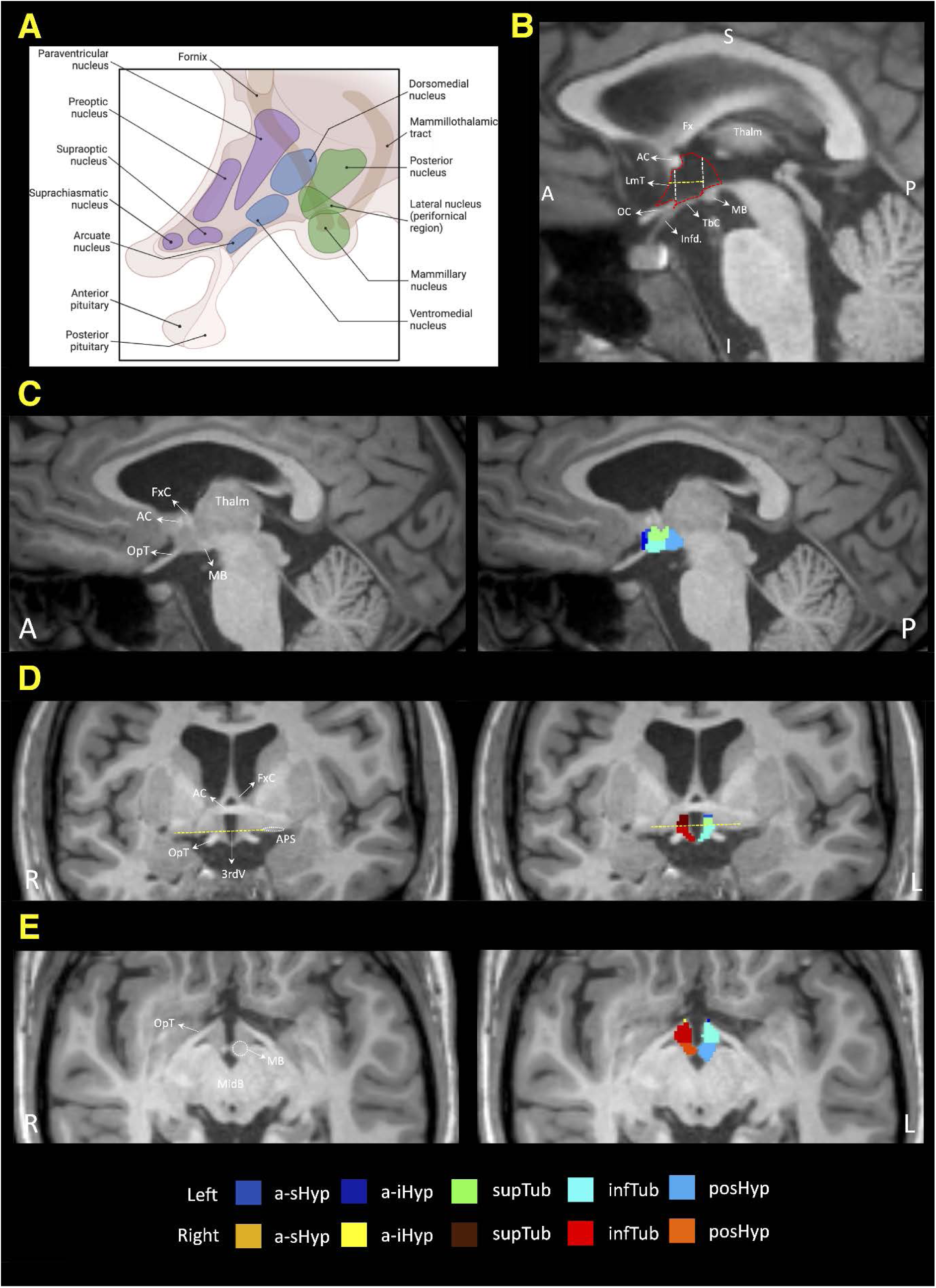
Hypothalamus and hypothalamic subunit anatomy on T1-weighted MRI. A: schematic showing the hypothalamic anatomy and conventional groupings of hypothalamic nuclei into an anterior (purple), a tubular (blue), and a posterior (green) group. The anatomical landmarks used to define the hypothalamus and its five subunits are labelled, in mid-sagittal (B), para-sagittal (C), coronal (D), and axial (E) planes. In B. the hypothalamus is highlighted by the red dotted outline. It is bounded antero-superiorly by the *anterior commissure* (AC), anteriorly by the *lamina terminalis* (LmT), and antero-inferiorly by the *optic chiasm* (OC); inferiorly by the *pituitary infundibulum* (infd), *tuber cinereum* (TbC), and the *mamillary body* (MB); posteriorly by the posterior extend of MB; superiorly by the *hypothalamic sulcus* and the *thalamus* (Thalm); and medially by the *third ventricle* (3rdV). At the *midbrain* (MidB) level, it is bounded laterally by the *optic tract* (OpT). The two white dotted lines shown in B mark the division of the three main hypothalamic groups. The anterior group is bounded anteriorly by the anterior most extend of AC, and posteriorly by the anterior most extend of the pituitary infundibulum. The tubular group is bounded anteriorly by the pituitary infundibulum, and posteriorly by the anterior most extend of the MB. The posterior group is bounded posteriorly by the posterior most extend of the MB. The yellow dotted line shown in B and D are at the level of the *anterior perforating substance* (APS, in D; a.k.a. the floor of *substantia innomianta*). It provides the anatomical landmark to split the anterior and tubular groups into respective superior and inferior subunits. The *post- commissural column of fornix* (FxC) terminates at the MB, thus, excluded from the hypothalamic segmentation. Other abbreviations: a-sHyp = anterior-superior subunit; a-iHyp = anterior-inferior subunit; supTub = superior tubular subunit; infTub = inferior tubular subunit; postHyp = posterior subunit; Fx = fornix; A = anterior; P = posterior; R = right; L = left; S = superior; I = inferior.

### Statistical Analysis

#### Accuracy and reliability of pituitary and hypothalamus segmentation

To assess the intra-rater test-retest reliability of the pituitary segmentations, Dr JYMY repeated tracing on a randomly chosen subset of 24 pituitary glands (14 Fontans, 10 controls), four months between the two tracing attempts. We reported both the intra-rater intra-class correlation coefficient (ICC) scores and the dice similarity coefficient (DSC) between the test-retest pituitary segmentations. The DSC score reflects the degree of spatial agreement between the two segmentation attempts, ranged between 0 (no overlap) to 1 (perfect overlap). Interpretations of both the ICC and DSC scores are as follows: poor (<0.40), fair (0.40 ≥ I, < 0.60), good (0.60 ≥, < 0.75), and excellent (≥ 0.75).

Separately, the same 24 cases were reviewed independently by the two study neuroradiologists (Dr SB and Dr WG), who were blinded to the case groupings, for pituitary gland segmentation accuracy.

Dr JYMY assessed all hypothalamus and subunit segmentation outputs, based on the anatomical landmarks used to define the hypothalamus boundaries and its five subunits. ^23^

#### Groupwise volumetric comparison

Generalized Linear Model (GLM) was carried out to compare the pituitary and hypothalamus volumes and other demographic and clinical variables between Fontan/Control groups and age groups (<18 and >18 years old). Variables examined included: demographic (sex, gestational age*, birth weight, * BMI, height percentile, clinical variables (time from Fontan to MRI (duration of Fontan circulation), age at first Fontan surgery), and Fontan surgical variables (number of cardiac operations prior to Fontan procedure, predominant ventricular morphology, type of Fontan procedure [AP, AP converted to ECC, ECC, LT, LT converted to ECC]. Two-way analysis of variance using Fontan/control and age groups was applied where appropriate. Multivariable regression with age as a continuous variable, by Fontan/Control groups by volumes was also carried out. Means and standard deviation was presented. Chi-squared test for categorical measures were presented as counts and percentages. Statistical analyses were carried out using SAS 9.4 (SAS Institute Inc., Cary, NC, USA.). p-values of <0.05 were considered significant. *Due to the low number of cases (<30%) with gestational age and birthweight data available, these were not included.

All volume metrics were normalized for total brain volume (TBV) by simple division, since the previous publication using the same Fontan cohort had demonstrated significantly smaller brain volumes compared to the age- and sex-matched healthy controls. ^8^ The normalized volumes were very small (i.e., in the order of 10^-4^), thus, we multiplied the values by 10^4^, to assist with readability and results interpretation.

## Results

Eighty-six cases from the Fontan registry were included, with eighty-six matched controls. Clinical and demographic data was available for eighty-five of the Fontan cases, with omission of data for one case. Only age and sex data were available for the published normative controls.

The mean age and sex demographics are comparable between the Fontan and control groups (Table 2). Height centiles and BMI were higher in the over 18-year-old group than the under 18-year-old group within the Fontan cohort (p = 0.08 and 0.003 respectively). Age (in days) at time of first cardiac surgery was greater in the over 18 group (260 days compared to 87 days) (p = 0.05).

**Table 2.**
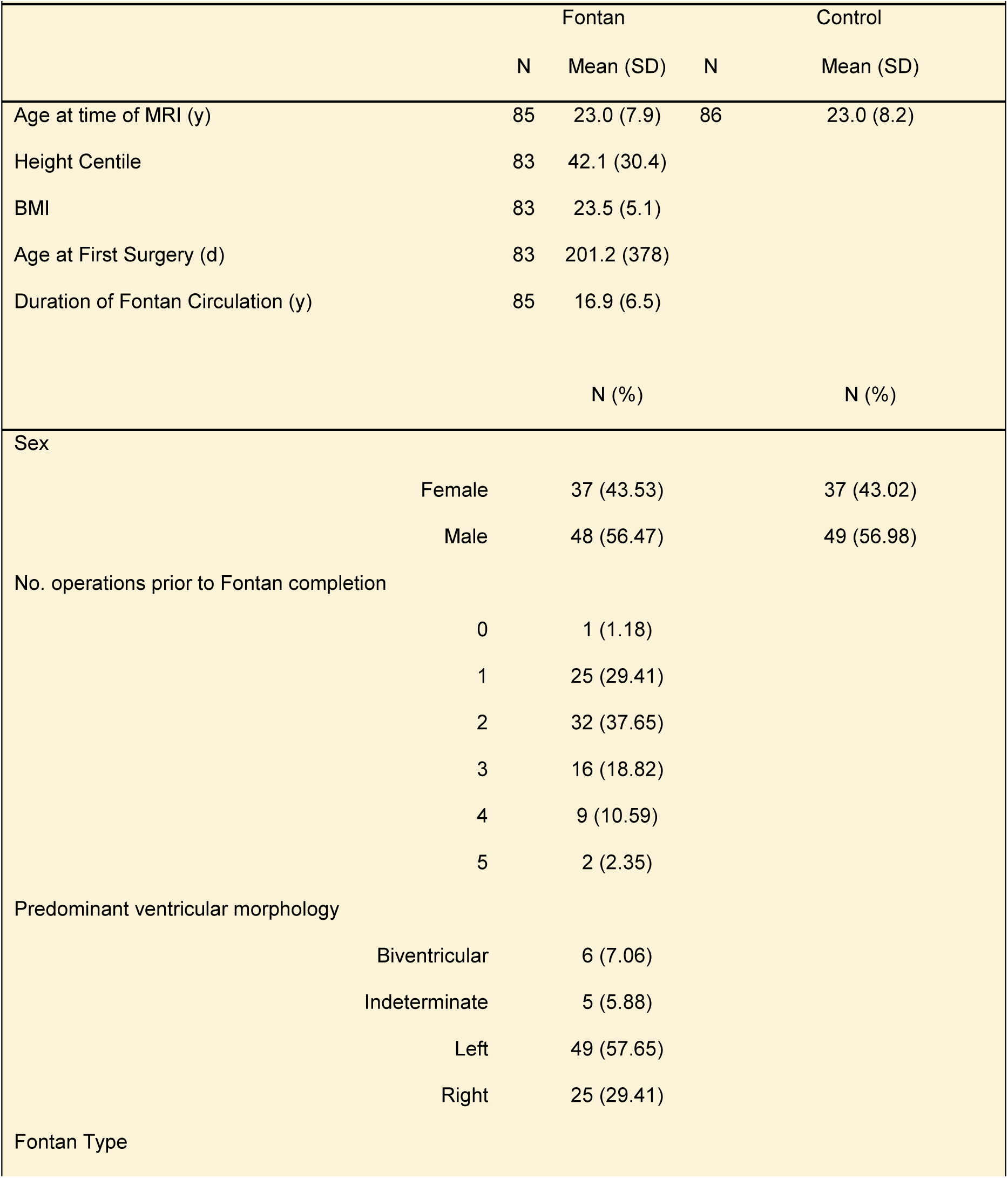
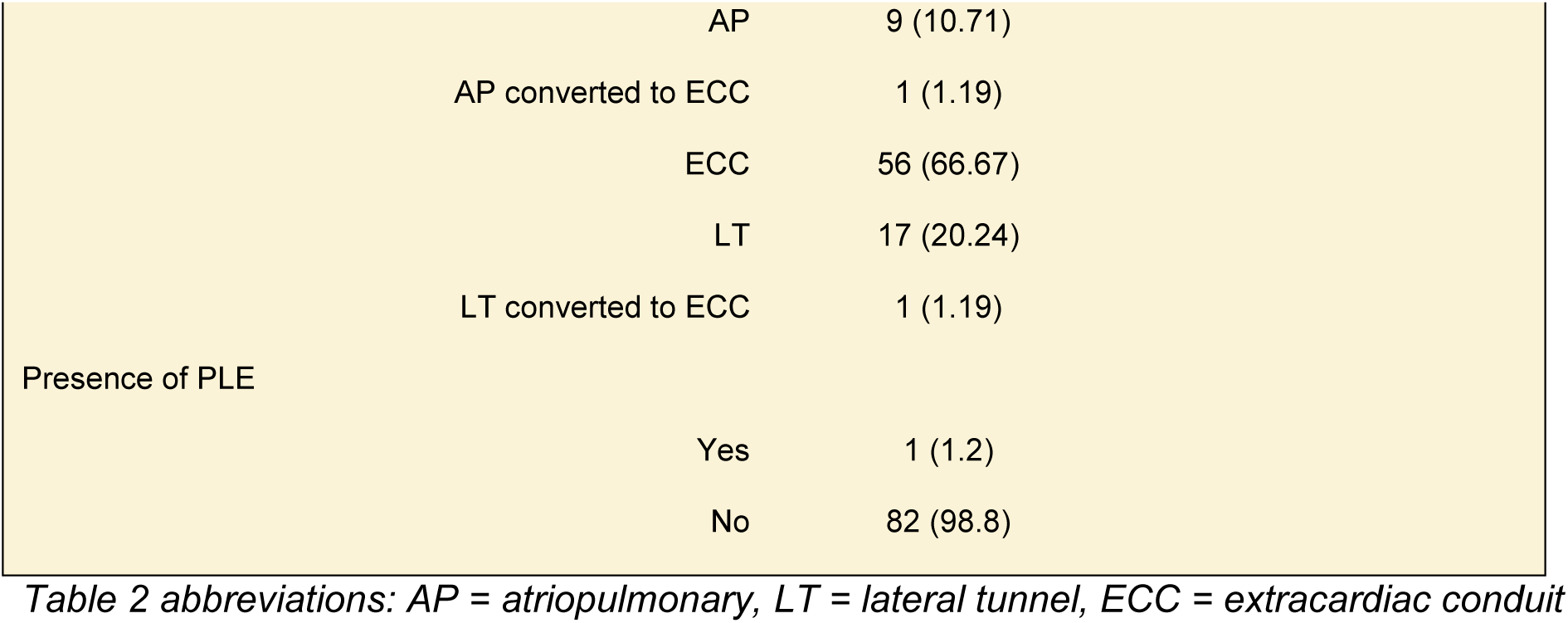
Clinical and Demographic Characteristics of Participants.

We excluded one Fontan case from pituitary volumetric analysis, who had an ectopic posterior pituitary gland, as a congenital abnormality. One control case had a partially empty sella. This case was included in the final analysis given it is considered a normal variant. ^24^

### Accuracy and Reliability of Pituitary and Hypothalamus Segmentation

The intra-rater ICC scores were 0.99 for all pituitary volumetrics. The DSC scores for all pituitary segmentations were (APV: median = 0.982, IQR = 0.978-0.988; PPV: median = 0.966, IQR = 0.948-0.977; TPV: median = 0.984, IQR = 0.980 -0.988).

The two study radiologists independently reviewed the twenty-four randomly selected sample cases and agreed all segmentations were anatomically accurate. The anatomical accuracy of all hypothalamus and subunit segmentation outputs were assessed and deemed satisfactory by the study neurosurgery research fellow (Dr JYMY).

Table 3 contains mean absolute and normalized volumetric measurements for each group. Please see supplementary material for age stratification. Table 4 demonstrates the mean differences in volumes between groups.

**Table 3.** Absolute and Normalized Pituitary and Hypothalamus Volumetric Measurements*. *Absolute gland volumes are measured in mm^3^, normalized gland volumes are a percentage of total brain volume scaled by a factor of 100.

|  | Fontan |  | Control |  |
| --- | --- | --- | --- | --- |
|  | N | Mean (SD) | N | Mean (SD) |
| TPV | 85 | 596.74 (134.09) | 86 | 526.94 (132.33) |
| APV | 64 | 480.14 (118.07) | 86 | 419.34 (114.8) |
| PPV | 64 | 103.02 (46.39) | 86 | 107.62 (45.56) |
| nTPV | 85 | 5.55 (1.54) | 86 | 4.38 (1.11) |
| nAPV | 64 | 4.31 (1.22) | 86 | 3.48 (0.97) |
| nPPV | 64 | 0.92 (0.43) | 86 | 0.89 (0.37) |
| RHV | 86 | 384.1 (47.9) | 86 | 388 (59.3) |
| R anterior-inferior | 86 | 13.95 (5.85) | 86 | 12.53 (7.05) |
| R anterior-superior | 86 | 21.51 (4.86) | 86 | 19.16 (6.76) |
| R posterior | 86 | 108.55 (21.82) | 86 | 119.48 (20.56) |
| R tubular inferior | 86 | 129.24 (15.27) | 86 | 126.37 (25.27) |
| R tubular superior | 86 | 110.84 (15.31) | 86 | 110.45 (20.03) |
| LHV | 86 | 392.0 (45.6) | 86 | 400.8 (50.1) |
| L anterior-inferior | 86 | 14.53 (5.11) | 86 | 13.8 (5.86) |
| L anterior-superior | 86 | 20.49 (4.05) | 86 | 20.34 (5.66) |
| L posterior | 86 | 106.58 (19.44) | 86 | 114.45 (17.59) |
| L tubular inferior | 86 | 140.08 (16.86) | 86 | 140.55 (20.92) |
| L tubular superior | 86 | 110.35 (15.34) | 86 | 111.62 (15.52) |
| nRHV | 86 | 3.51 (0.36) | 86 | 3.21 (0.49) |
| nR anterior-inferior | 86 | 0.13 (0.05) | 86 | 0.1 (0.06) |
| nR anterior-superior | 86 | 0.2 (0.04) | 86 | 0.16 (0.05) |
| nR posterior | 86 | 0.99 (0.18) | 86 | 0.99 (0.18) |
| nR tubular inferior | 86 | 1.18 (0.13) | 86 | 1.04 (0.20) |
| nR tubular superior | 86 | 1.02 (0.13) | 86 | 0.92 (0.17) |
| nLHV | 86 | 3.59 (0.38) | 86 | 3.32 (0.40) |
| nL anterior-inferior | 86 | 0.13 (0.05) | 86 | 0.11 (0.05) |
| nL anterior-superior | 86 | 0.19 (0.04) | 86 | 0.17 (0.05) |
| nL posterior | 86 | 0.97 (0.16) | 86 | 0.95 (0.15) |
| nL tubular inferior | 86 | 1.28 (0.15) | 86 | 1.16 (0.16) |
| nL tubular superior | 86 | 1.01 (0.14) | 86 | 0.93 (0.13) |

**Table 4.** Mean Differences in Normalized Pituitary and Hypothalamus Volumes Between Groups.

|  | Group Difference (Fontan - Control) | 95% CI | p |
| --- | --- | --- | --- |
|  | Estimate (SE) |  |  |
| nTPV | 1.17 (0.20) | 0.77 - 1.57 | **<.0001** |
| nAPV | 0.81 (0.17) | 0.47 - 1.15 | **<.0001** |
| nPPV | 0.02 (0.06) | -0.25 | 0.72 |
| nLHV | 0.27 (0.06) | 0.15 - 0.39 | **<.0001** |
| nL anterior-inferior | 0.02 (0.01) | 0.01 - 0.03 | **0.009** |
| nL anterior-superior | 0.02 (0.01) | 0.01 - 0.03 | **0.002** |
| nL posterior | 0.03 (0.02) | -0.02 - 0.07 | 0.28 |
| nL tubular inferior | 0.12 (0.02) | 0.07 - 0.17 | **<.0001** |
| nL tubular superior | 0.09 (0.02) | 0.05 - 0.13 | **<.0001** |
| nRHV | 0.30 (0.07) | 0.17 - 0.43 | **<.0001** |
| nR anterior-inferior | 0.02 (0.01) | 0.01 - 0.04 | **0.0073** |
| nR anterior-superior | 0.04 (0.01) | 0.02 - 0.05 | **<.0001** |
| nR posterior | 0.0003 (0.03) | -0.05 - 0.05 | 0.99 |
| nR tubular inferior | 0.14 (0.03) | 0.09 - 0.19 | **<.0001** |
| nR tubular superior | 0.1 (0.02) | 0.05 - 0.15 | **<.0001** |

Absolute TPV, APV and PPV were not significantly different between the Fontan and Control groups. When normalized for total brain volume, the mean nTPV and nAPV were significantly higher in the Fontan group compared to the controls (*p* <0.0001 for both). The mean difference between the groups for nTPV was 1.17 units (gland volume as a percentage of total brain volume x 100), and for nAPV was 0.81 units. There was no significant difference in mean nPPV between groups (*p* = 0.5285), which suggested the increased TPV in the Fontan group was driven by enlarged APV (Figure 3).

**Figure 3:**
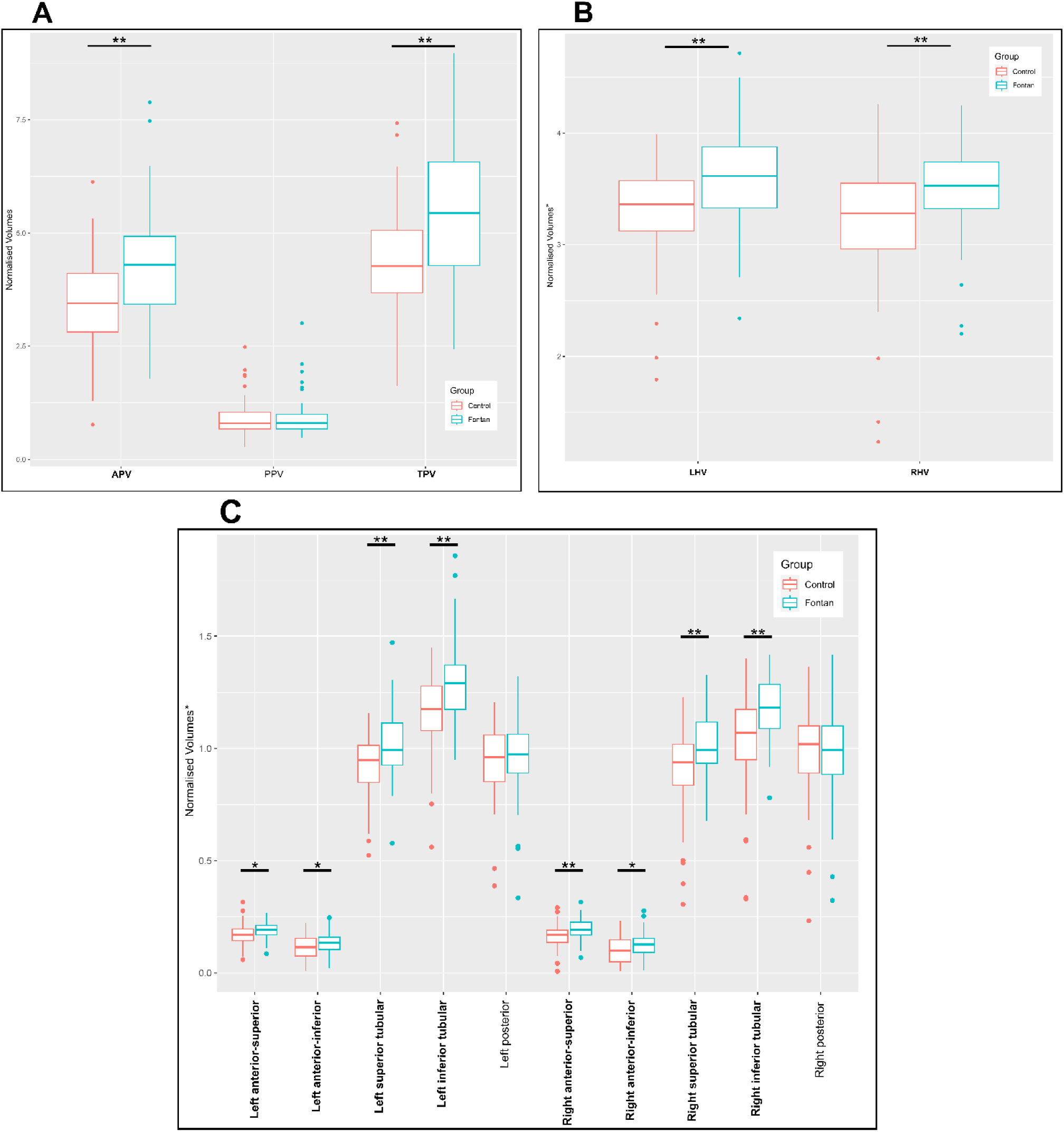
Group comparisons of normalised pituitary (A), bilateral hypothalamus (B) and hypothalamus subunit (C) volumes. Note the normalised pituitary and hypothalamus volumetric values are a percentage of total brain volume scaled by a factor of 100. Differences between groups for anterior and total pituitary, bilateral hypothalamus, and bilateral anterior and tubular subunit volumes were significant (bolded). *p<0.001, **p<0.0001. The differences between posterior pituitary and posterior hypothalamus subunit volumes were not significant.

Normalized bilateral hypothalamus volumes (nLHV and nRHV) were both significantly larger in Fontan patients compared to controls, with mean difference 0.28 and 0.30 units respectively (*p* values <0.0001). Normalized bilateral anterior and tubular hypothalamic subunits were also significantly larger in Fontan patients than controls (*p* <0.0087 and <0.0001, respectively). The posterior hypothalamic subunit volumes were not significantly different between groups (*p* = 0.9767 [right)]and *p* = 0.2703 [left)] (Figure 3).

Age stratification above and below 18 years did not have a significant effect on any of the mean measured volumes. Multivariable regression analysis with age as a continuous variable demonstrated a significant positive association between nTPV and increasing age (nTPV increases by 0.03 units per year of age, *p* = 0.03). The interaction term for nTPV was not significant (*p* = 0.82), suggesting that the association between nTPV and age did not differ significantly between the Fontan and Control groups. There was a significant positive association between both nAPV and nPPV with age (*p* = 0.002 and 0.004). The interaction term for nAPV was significant, *p* = 0.0046, suggesting the increase in nAPV with age is greater in the Fontan group than the Control group; this is illustrated in Figure 4. The interaction term for nPPV was not significant. There was no significant association between nRHV or nLHV with age (*p* = 0.46 and 0.11).

**Figure 4:**
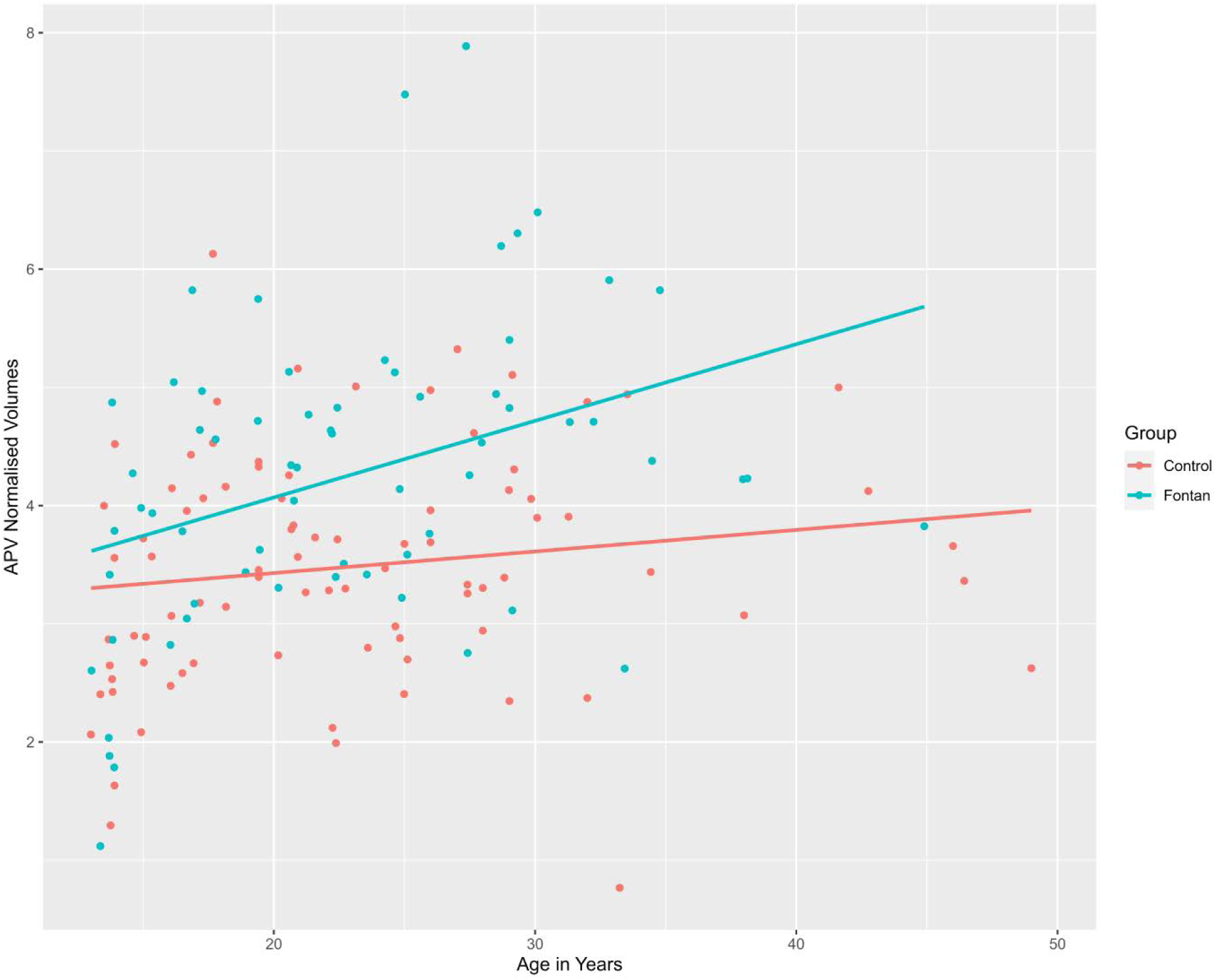
Group comparison of the relationship between age (years) and normalized anterior pituitary volumes, with lines of best fit. Note the normalized pituitary volumes are a percentage of total brain volume scaled by a factor of 100.

There are significant positive associations between nTPV and nAPV with increasing time between Fontan and MRI acquisition (i.e., duration of the Fontan circulation), (*p* = 0.04 and 0.002). There are two apparent peaks at approximately 14 years and 25 years post-Fontan when this data is presented in graphical format Please refer to the supplementary material.

The height centile had a significant positive association with nAPV (*p* = 0.0163). There were no significant associations with the remaining clinical and surgical variables investigated: sex, age at first surgery, BMI, number of cardiac surgeries prior to Fontan, predominant ventricular morphology, and Fontan type (*p*-values ranged 0.1 to 0.9; see supplementary material Tables 5 - 9). Protein-losing enteropathy was only present in one case, precluding statistical analysis.

## Discussion

We have demonstrated that normalized anterior pituitary and anterior and tubular hypothalamic subunit volumes were increased in those with a Fontan circulation compared to controls, and anterior pituitary volumes in particular were more significantly increased with age. These findings support the hypothesis that the HPA and the hypothalamic-pituitary portal venous system are affected by the Fontan circulation.

The enlargement of the anterior pituitary independent of the posterior pituitary is in keeping with the physiology of the pituitary vascular supply; the anterior pituitary gland is supplied by the portal circulation (blood descends from the hypothalamus along the infundibulum in the hypophyseal portal venous plexus), whereas the posterior pituitary is supplied by arterial branches from the systemic circulation (branches from the internal carotid artery). ^25^ Enlargement of the anterior pituitary and anterior and tubular hypothalamic subunits could be either due to venous congestion or potentially if there is endocrine feedback or “hyperstimulation of the hypothalamic-pituitary axis”. The nAPV increases with increasing height centile; it is likely the two are related, as the anterior pituitary releases growth hormone. Future studies including collection of comprehensive hormonal data and circulatory data that can directly or indirectly infer portal venous pressure in the hypothalamic-pituitary system are required to disentangle the potential pathophysiological mechanisms behind our study observation.

The finding of larger normalized hypothalamic nuclei volumes in Fontan participants supports the effect of hyperstimulation of the hypothalamic pituitary axis due to downstream hormone deficiency, or from the “upstream” congestion of the portal system; this study cannot delineate. Increased bilateral normalized anterior and tubular subunit group volumes were noted in Fontan participants. The arcuate nucleus, located in the inferior tubular subunit, secretes various hormones into the hypothalamic-pituitary portal venous system to act on the anterior pituitary gland. These hormones include growth hormone releasing hormone (GHRH), thyrotropin-releasing hormone (TRH) and gonadotrophin-releasing hormone, among others. ^26^

GHRH triggers growth hormone (GH) release by the anterior pituitary gland. Alteration in this mechanism may be implicated in the high incidence of short stature seen in Fontan populations.^3,6^ TRH triggers thyroid-stimulating hormone (TSH) production by the anterior pituitary gland; this system may be involved in the high incidence of hypothyroidism as well as short stature in Fontan populations.^3,4,6^

Gonadotropin-releasing hormone (GnRH) neurons are found in the medial preoptic nucleus (in the anterior superior subunit) and in the arcuate nucleus. These trigger follicular stimulating hormone (FSH) and luteinizing hormone (LH) production by the anterior pituitary gland via the portal circulation. ^27^ Subsequent deficiency in estrogen and testosterone production may be a potential explanation for the high frequency of hypogonadism seen in Fontan populations.^5,6^

A significant positive association was demonstrated between nTPV and age in all subjects, with no difference in this relationship between Fontan and Control groups. This may suggest a “normal” age related phenomenon. However, when total pituitary was divided into anterior and posterior glands, the increase in nAPV with age was greater in the Fontan group compared to the controls. This suggests a possible element of congestion in Fontan circulations contributing to the anterior pituitary volume increase over time.

Both the normalized TPVs and APVs were seen to increase with time since Fontan with two apparent peaks at 14 years and 25 years post-Fontan. Note the mean age of Fontan completion was 6.2 years. A possible explanation for the first peak may be onset of (delayed) puberty. ^28^ The second peak at 25 years post-Fontan aligns with published evidence that pituitary heights peak in the age group between 20 -29 in a normal population, however the mechanism for this is unclear.^29^

## Strengths and Limitations

Observations from Muneuchi (2018) in 40 children post Fontan surgery showed significantly larger pituitary volumes on brain MRI compared to a control population (nearly 80% larger) and correlated this to higher central venous pressures. ^7^ The reported pituitary volumes were estimates based on dimensional measurements (height, depth and width measured on T1- weighted sagittal and coronal images), and anterior or posterior gland measurements were not assessed separately. To date this study’s findings had not yet been replicated.

Access to data from the ANZ Fontan Registry for our study allowed analysis of a larger Fontan population than previously reported. The volumetric analyses performed in our study utilized both expert-based manual segmentation and deep-learning based automated image segmentation techniques, the accuracy of which was independently validated by anatomical experts. Our adopted approach enables region specific image segmentations (e.g., the ability to compute the APV and PPV, and hypothalamus subunit volumes), and the 3D nature of these segmentations are preferred over volumes estimated based on 2D dimensional measurements from MRI in standard orthogonal planes. To our best knowledge, this is the first study that investigated the hypothalamus and hypothalamus subunit volumetrics in the Fontan patients.

The current study is an exploratory retrospective analysis utilizing a clinical and MRI dataset from an existing ANZ Fontan Registry study. This meant the exclusion criteria may not necessarily align with exclusion criteria we may have proposed if recruiting participants prospectively. Patients with severe intellectual disability and/or genetic syndromes with cognitive impairment were excluded from the studies, thus the participants studied may not reflect the worst-functioning Fontan population. ^8^ Retrospective data collection also meant that a small number of cases had incomplete clinical and demographic datasets. Lack of comprehensive hormonal data and circulatory data precluded us to make direct statements about the status of endocrine function and HPA portal venous pressure of the Fontan participants, at the time when the study data was being collected.

As with all multi-site MRI studies, a degree of heterogeneity due to variation in scanner hardware can be an issue. All of the Fontan participant MRIs were acquired on hardware from a single vendor and with tightly controlled acquisition parameters, which helped reduce variance. We utilized open-source normative control MRI data due to lack of recruitment of study specific normative controls. The normative control data from the ABIDE and PING databases was acquired by different centers on hardware from varying vendors. Furthermore, we have assumed the normative controls described in the ABIDE and PING databases are without congenital heart disease; this is only explicitly confirmed in a few of the ABIDE constituent cohorts, and not specified for the PING participants. ^9,10,30^

Future prospective studies in the current or separate Fontan cohort with co-recruitment of study-specific healthy controls, and collection of comprehensive hormonal and circulatory data may be able to shed light on exact pathophysiological mechanism accounting for the observed enlargement of anterior pituitary and hypothalamus volumes in this study.

## Conclusions

This study from the ANZ Fontan registry has shown that adolescents and adults with a Fontan circulation have increased anterior pituitary volumes, and larger hypothalamus volumes (specifically anterior and tubular hypothalamic subunit groups). This supports the hypothesis that the hypothalamic-pituitary portal venous system is also affected by duration of Fontan circulation, therefore increasing the risk of progressive HPA dysfunction in these patients over time. Our findings suggest that monitoring growth, puberty and pituitary function in children and adolescents following Fontan surgery is warranted, and further prospective studies may consider correlating cardiac, venous and HPA evaluations with clinical outcomes and growth.

## Data Availability

The ANZ Fontan Registry data used in this study was acquired from clinical patients. They were not made openly available due to ethics issue of handling clinical data.

The MRI data used in this study was acquired from clinical patients.

https://fcon_1000.projects.nitrc.org/indi/abide/

http://pingstudy.ucsd.edu/

## Author Contribution

**WG**: Contributed to study conceptualization, methodology, validation, major role in writing the original draft and project co-ordination. **JYMY**: Contributed to study conceptualization and imaging methodology, projection co-ordination, computation resources, major role imaging data curation and processing, assisted with drafting original manuscript and review. **TG**: Contributed to study conceptualization and review of final manuscript. **SB**: Contributed to validation, review of final manuscript. **AI**: Contributed to study conceptualization and review of final manuscript. **JC**: Contributed to imaging methodology design, major role in data curation, processing and analysis. **DYH**: Major role in formal statistical analysis, and figure preparation. **RC**: Contributed to review of final manuscript. **CV**: Contributed to data curation and review of final manuscript. **CJ**: Contributed to study conceptualization, assisted with drafting original manuscript and review of final manuscript, provided supervision for the project.

## Acknowledgements

JYMY and JC acknowledge the support of the Royal Children’s Hospital, Murdoch Children’s Research Institute, The University of Melbourne Department of Paediatrics, and the Victorian Government’s Operational Infrastructure Support Program.

The normative control MRI data used in the preparation of this article were obtained from the ABIDE I and ABIDE II (Autism Brain Imaging Data Exchange) data sets and the PING (Pediatric Imaging, Neurocognition and Genetics Study) database. Data collection and sharing for this project was funded by PING (National Institutes of Health [NIH]: RC2DA029475). PING is funded by the National Institute on Drug Abuse and the Eunice Kennedy Shriver National Institute of Child Health & Human Development (NIH: RC2DA029475). PING data are disseminated by the PING Coordinating Center at the Center for Human Development, University of California, San Diego. As such, the investigators within PING contributed to the design and implementation of PING and provided data but did not participate in analysis or writing of this report. A complete listing of PING investigators can be found at the PING database website. The authors would also like to thank A. Di Martino, M. P. Milham, and all involved in the collection and aggregation of the ABIDE I and ABIDE II data sets. Figure 2A was created with BioRender.com.

