## Supplemental Information for "Segmental MRI Pituitary and Hypothalamus Volumes post Fontan: An analysis of the Australian and New Zealand Fontan Registry"

### SUPPLEMENTAL MATERIAL:

#### Supplemental Methods:

*Brain MRI Acquisition*

Brain MRI data was acquired using Siemens 3Tesla MRI scanners: RCH (TrioTim), RPAH (Skyra), CHW (Verio), and NZ (Skyra). The sequences used for segmentation included 3D T1-weighted imaging and 2D T2-weighted spin echo imaging reconstructed in three standard orthogonal planes. T2-weighted Fluid Attenuated Inversion Recovery (FLAIR) sequence, diffusion-weighted imaging (DWI), and Susceptibility-Weighted Imaging (SWI) were acquired for the Verrall study, but not used for this retrospective analysis.
The 3D T1-weighted images were used for brain volumetric analysis. These were acquired using the 3D Magnetization Prepared Rapid Gradient Echo (MPRAGE) sequence at the RCH, RPAH and CHW sites (in-plane voxel resolution ~0.45 to 0.94 mm; slice thickness = 0.9 mm; TR = 1680 to 2530 ms; TE = 1.77 to 2.45 ms; FOV ~ 230 x 230 mm; Flip angle = 7 to 9 degrees); and the 3D Magnetization Prepared 2 Rapid Gradient Echo (MP2RAGE) sequence for the NZ site (1.0mm isotropic voxel resolution; TR = 5000msec; TE = 2.50msec; FOV = 256 x 256mm; Flip angle = 0 degree).

Please refer to the ABIDE I and II and PING data repository websites and publications for acquisition parameter details, and the criteria used for the recruited healthy control subjects.

*Pituitary Gland Segmentation Protocol*

Pituitary segmentation described by Farrow et al., 2020:

"*Pituitary gland tracing is performed in the coronal planes, given it provides the best visualization of the pituitary gland (Garner et al., 2005; Lorenzetti et al., 2009). The borders of pituitary gland are defined by the diaphragm sellae superiorly, the sphenoid sinus inferiorly, and the cavernous sinuses bilaterally. The infundibular stalk is excluded from the tracing (Ganella et al., 2015; Whittle et al., 2012). The whole pituitary gland tracing is then subsequently split into anterior and posterior lobes. The difference between the darker (i.e. T1-weighted isointense) anterior lobe and T1-weighted hyperintense posterior lobe is typically easily identifiable. This division is first identified in the coronal plane, but for cases where the divide is not parallel with a coronal slice, further editing is performed in the sagittal plane, where the boundary could be clarified*." (Figure 1).

Note in this protocol, the anterior pituitary gland tracing includes both the *pars distalis* and *pars intermedia*, but excludes the *pars tuberalis* which forms part of infundibulum/pituitary stalk.

SUPPLEMENTAL TABLES:

**Table 2a: Clinical and Demographic Characteristics of Participants (with age group stratification)**

| | **Fontan** | | | | | | | **Control** | | | | | | |
|---|---|---|---|---|---|---|---|---|---|---|---|---|---|---|
| | Total | | Age < 18 y | | Age ≥ 18 y | | | Total | | Age < 18 y | | Age ≥ 18 y | | |
| | N | Mean (SD) | N | Mean (SD) | N | Mean (SD) | p | N | Mean (SD) | N | Mean (SD) | N | Mean (SD) | p |
| **Age_yr** | 85 | 23.0 (7.9) | 28 | 15.4 (1.5) | 57 | 26.8 (6.9) | <.0001 | 86 | 23.0 (8.2) | 29 | 15.3 (1.5) | 57 | 27 (7.3) | <.0001 |
| **Ht_centile** | 83 | 42.1 (30.4) | 28 | 33.9 (28.9) | 55 | 46.3 (30.5) | 0.0783 | | | | | | | |
| **BMI** | 83 | 23.5 (5.1) | 28 | 21.3 (4.9) | 55 | 24.7 (4.8) | 0.0034 | | | | | | | |
| **Age1st_surgery_days** | 83 | 201.2 (378) | 28 | 86.8 (133) | 55 | 260 (445) | 0.0483 | | | | | | | |
| **Time_Fontan_MRI_yr** | 85 | 16.9 (6.5) | 28 | 10.4 (1.8) | 57 | 20.1 (5.6) | <.0001 | | | | | | | |
| **Age_at_Fontan_yrs** | 84 | 6.2 (4.6) | 28 | 4.9 (1.0) | 56 | 6.8 (5.5) | 0.0778 | | | | | | | |

### Table 2b: Clinical and Demographic Characteristics of Participants (with age group stratification)

| | | Fontan | | | | Control | | | |
|---|---|---|---|---|---|---|---|---|---|
| | | Total<br>N (%) | Age < 18 y<br>N (%) | Age ≥ 18 y<br>N (%) | p | Total<br>N (%) | Age < 18 y<br>N (%) | Age ≥ 18 y<br>N (%) | p |
| | Total | 85 (100) | 28 (32.94) | 57 (67.06) | | 86 (100) | 29 (33.72) | 57 (66.28) | |
| **Sex** | F | 37 (43.53) | 9 (32.14) | 28 (49.12) | 0.1667 | 37 (43.02) | 9 (31.03) | 28 (49.12) | 0.1664 |
| | M | 48 (56.47) | 19 (67.86) | 29 (50.88) | | 49 (56.98) | 20 (68.97) | 29 (50.88) | |
| **No_operations_prior_Fontan** | 0 | 1 (1.18) | 0 (0) | 1 (1.75) | 0.1107 | | | | |
| | 1 | 25 (29.41) | 4 (14.29) | 21 (36.84) | | | | | |
| | 2 | 32 (37.65) | 16 (57.14) | 16 (28.07) | | | | | |
| | 3 | 16 (18.82) | 5 (17.86) | 11 (19.3) | | | | | |
| | 4 | 9 (10.59) | 3 (10.71) | 6 (10.53) | | | | | |
| | 5 | 2 (2.35) | 0 (0) | 2 (3.51) | | | | | |
| **Predominant_ventricular_morpholo** | Biventricular | 6 (7.06) | 3 (10.71) | 3 (5.26) | 0.7065 | | | | |
| | Indeterminate | 5 (5.88) | 1 (3.57) | 4 (7.02) | | | | | |
| | Left | 49 (57.65) | 15 (53.57) | 34 (59.65) | | | | | |
| | Right | 25 (29.41) | 9 (32.14) | 16 (28.07) | | | | | |
| **Fontan_Type Age_gp** | AP | 9 (10.71) | 0 (0) | 9 (16.07) | 0.0003 | | | | |
| **(this p-value is not meaningful)** | APconvertedtoECC | 1 (1.19) | 0 (0) | 1 (1.79) | | | | | |
| | ECC | 56 (66.67) | 28 (100) | 28 (50) | | | | | |
| | LT | 17 (20.24) | 0 (0) | 17 (30.36) | | | | | |
| | LTconvertedtoECC | 1 (1.19) | 0 (0) | 1 (1.79) | | | | | |
| **Presence_PLE Age_gp** | Yes | 1 (1.2) | 1 (3.7) | 0 (0) | 0.3253 | | | | |
| | No | 82 (98.8) | 26 (96.3) | 56 (100) | | | | | |

**Table 3 (complete with age stratification):** ***Absolute and Normalized Pituitary and Hypothalamus Volumetric Measurements***

| | *Fontan* | | | | | | *Control* | | | | | |
|---|---|---|---|---|---|---|---|---|---|---|---|---|
| | Total | | Age < 18 y | | Age ≥ 18 y | | Total | | Age < 18 y | | Age ≥ 18 y | |
| | N | Mean (SD) | N | Mean (SD) | N | Mean (SD) | N | Mean (SD) | N | Mean (SD) | N | Mean (SD) |
| *TPV* | 85 | 596.74 (134.09) | 28 | 561.82 (142.91) | 57 | 613.89 (127.34) | 86 | 526.94 (132.33) | 29 | 499.03 (149.8) | 57 | 541.14 (121.45) |
| *APV* | 64 | 480.14 (118.07) | 20 | 424.5 (117.54) | 44 | 505.43 (110.57) | 86 | 419.34 (114.8) | 29 | 393.79 (132.95) | 57 | 432.33 (103.24) |
| *PPV* | 64 | 103.02 (46.39) | 20 | 93.75 (38.39) | 44 | 107.23 (49.43) | 86 | 107.62 (45.56) | 29 | 105.24 (39.36) | 57 | 108.82 (48.7) |
| *TPV_norm* | 85 | 5.55 (1.54) | 28 | 5.22 (1.73) | 57 | 5.72 (1.42) | 86 | 4.38 (1.11) | 29 | 4.11 (1.24) | 57 | 4.51 (1.02) |
| *APV_norm* | 64 | 4.31 (1.22) | 20 | 3.66 (1.14) | 44 | 4.60 (1.16) | 86 | 3.48 (0.97) | 29 | 3.24 (1.07) | 57 | 3.61 (0.90) |
| *PPV_norm* | 64 | 0.92 (0.43) | 20 | 0.79 (0.30) | 44 | 0.97 (0.46) | 86 | 0.89 (0.37) | 29 | 0.87 (0.34) | 57 | 0.09 (0.38) |
| *RHV* | 86 | 384.1 (47.9) | 29 | 378.2 (56.5) | 57 | 387.1 (43.1) | 86 | 388 (59.3) | 29 | 386.6 (64.6) | 57 | 388.7 (57.0) |
| *R_anterior_inferior_hypo* | 86 | 13.95 (5.85) | 29 | 13.03 (5.88) | 57 | 14.42 (5.84) | 86 | 12.53 (7.05) | 29 | 11.07 (6.18) | 57 | 13.28 (7.4) |
| *R_anterior_superior_hypo* | 86 | 21.51 (4.86) | 29 | 21.41 (5.25) | 57 | 21.56 (4.69) | 86 | 19.16 (6.76) | 29 | 17.93 (6.46) | 57 | 19.79 (6.88) |
| *R_posterior_hypo* | 86 | 108.55 (21.82) | 29 | 105.48 (27.43) | 57 | 110.11 (18.42) | 86 | 119.48 (20.56) | 29 | 126.31 (20.55) | 57 | 116 (19.84) |
| *R_tubular_inferior_hypo* | 86 | 129.24 (15.27) | 29 | 126.76 (15.09) | 57 | 130.51 (15.34) | 86 | 126.37 (25.27) | 29 | 124.24 (28.09) | 57 | 127.46 (23.89) |
| *R_tubular_superior_hypo* | 86 | 110.84 (15.31) | 29 | 111.55 (18.89) | 57 | 110.47 (13.31) | 86 | 110.45 (20.03) | 29 | 107.03 (25.77) | 57 | 112.19 (16.37) |
| *LHV* | 86 | 392.0 (45.6) | 29 | 386.1 (57.3) | 57 | 395 (38.6) | 86 | 400.8 (50.1) | 29 | 394.6 (49.4) | 57 | 403.9 (50.6) |
| *L_anterior_inferior_hypo* | 86 | 14.53 (5.11) | 29 | 11.9 (5.17) | 57 | 15.88 (4.56) | 86 | 13.8 (5.86) | 29 | 11.1 (5.86) | 57 | 15.18 (5.41) |
| *L_anterior_superior_hypo* | 86 | 20.49 (4.05) | 29 | 19.79 (4.11) | 57 | 20.84 (4.02) | 86 | 20.34 (5.66) | 29 | 19.1 (5.60) | 57 | 20.96 (5.64) |
| *L_posterior_hypo* | 86 | 106.58 (19.44) | 29 | 105.9 (26.38) | 57 | 106.93 (15.0) | 86 | 114.45 (17.59) | 29 | 115.59 (17.26) | 57 | 113.88 (17.89) |
| *L_tubular_inferior_hypo* | 86 | 140.08 (16.86) | 29 | 141.24 (18.23) | 57 | 139.49 (16.26) | 86 | 140.55 (20.92) | 29 | 138.83 (18.65) | 57 | 141.42 (22.1) |

| | | | | | | | | | | | | |
|---|---|---|---|---|---|---|---|---|---|---|---|---|
| *L_tubular_superior_hypo* | 86 | 110.35 (15.34) | 29 | 107.31 (18.73) | 57 | 111.89 (13.22) | 86 | 111.62 (15.52) | 29 | 110 (14.82) | 57 | 112.44 (15.92) |
| *RHV_norm* | 86 | 3.51 (0.36) | 29 | 3.41 (0.42) | 57 | 3.57 (0.32) | 86 | 3.21 (0.49) | 29 | 3.17 (0.53) | 57 | 3.24 (0.47) |
| *R_ant_inf_norm* | 86 | 0.13 (0.05) | 29 | 0.12 (0.05) | 57 | 0.13 (0.05) | 86 | 0.1 (0.06) | 29 | 0.09 (0.05) | 57 | 0.11 (0.06) |
| *R_ant_sup_norm* | 86 | 0.2 (0.04) | 29 | 0.19 (0.05) | 57 | 0.20 (0.04) | 86 | 0.16 (0.05) | 29 | 0.15 (0.05) | 57 | 0.16 (0.06) |
| *R_post_norm* | 86 | 0.99 (0.18) | 29 | 0.95 (0.23) | 57 | 1.01 (0.15) | 86 | 0.99 (0.18) | 29 | 1.04 (0.18) | 57 | 0.97 (0.17) |
| *R_tub_inf_norm* | 86 | 1.18 (0.13) | 29 | 1.15 (0.15) | 57 | 1.2 (0.11) | 86 | 1.04 (0.20) | 29 | 1.02 (0.23) | 57 | 1.06 (0.18) |
| *R_tub_sup_norm* | 86 | 1.02 (0.13) | 29 | 1.01 (0.14) | 57 | 1.02 (0.13) | 86 | 0.92 (0.17) | 29 | 0.88 (0.21) | 57 | 0.94 (0.14) |
| *LHV_norm* | 86 | 3.59 (0.38) | 29 | 3.49 (0.47) | 57 | 3.64 (0.31) | 86 | 3.32 (0.40) | 29 | 3.23 (0.37) | 57 | 3.36 (0.41) |
| *L_ant_inf_norm* | 86 | 0.13 (0.05) | 29 | 0.11 (0.04) | 57 | 0.15 (0.04) | 86 | 0.11 (0.05) | 29 | 0.09 (0.05) | 57 | 0.13 (0.05) |
| *L_ant_sup_norm* | 86 | 0.19 (0.04) | 29 | 0.18 (0.04) | 57 | 0.19 (0.04) | 86 | 0.17 (0.05) | 29 | 0.16 (0.04) | 57 | 0.17 (0.05) |
| *L_post_norm* | 86 | 0.97 (0.16) | 29 | 0.95 (0.2) | 57 | 0.99 (0.13) | 86 | 0.95 (0.15) | 29 | 0.95 (0.13) | 57 | 0.95 (0.15) |
| *L_tub_inf_norm* | 86 | 1.28 (0.15) | 29 | 1.28 (0.19) | 57 | 1.29 (0.13) | 86 | 1.16 (0.16) | 29 | 1.14 (0.14) | 57 | 1.18 (0.17) |
| *L_tub_sup_norm* | 86 | 1.01 (0.14) | 29 | 0.97 (0.17) | 57 | 1.03 (0.12) | 86 | 0.93 (0.13) | 29 | 0.9 (0.12) | 57 | 0.94 (0.13) |

**Table 4a:** ***Mean Differences in Pituitary and Hypothalamus Volumes Between Groups*** **(with interaction term for age)**

| | Group | Estimate (SE) | 95% CI | p for Group | Age_gp | Estimate (SE) | 95% CI | p for Age | p for Group * Age |
|---|---|---|---|---|---|---|---|---|---|
| **TPV** | Fontan | 62.8 (35) | -6.3 - 131.9 | 0.0746 | 18<= | 42.1 (30.1) | -17.4 - 101.6 | 0.1641 | 0.8164 |
| | Control | 0 | 0 | | <18 | 0 | 0 | | |
| **APV** | Fontan | 30.7 (33) | -34.5 - 95.9 | 0.3534 | 18<= | 38.5 (25.9) | -12.6 - 89.7 | 0.1386 | 0.292 |
| | Control | 0 | 0 | | <18 | 0 | 0 | | |
| **PPV** | Fontan | -11.5 (13.4) | -37.9 - 14.9 | 0.3917 | 18<= | 3.58 (10.5) | -17.2 - 24.3 | 0.7334 | 0.5437 |
| | Control | 0 | 0 | | <18 | 0 | 0 | | |
| **TPV_norm** | Fontan | 1.12 (0.35) | 0.42 - 1.81 | 0.0018 | 18<= | 0.41 (0.3) | -0.19 - 1.01 | 0.1803 | 0.838 |
| | Control | 0 | 0 | | <18 | 0 | 0 | | |
| **APV_norm** | Fontan | 0.43 (0.3) | -0.17 - 1.03 | 0.1612 | 18<= | 0.37 (0.24) | -0.1 - 0.84 | 0.1197 | 0.1289 |
| | Control | 0 | 0 | | <18 | 0 | 0 | | |
| **PPV_norm** | Fontan | -0.08 (0.11) | -0.3 - 0.15 | 0.4991 | 18<= | 0.04 (0.09) | -0.14 - 0.21 | 0.6932 | 0.2871 |
| | Control | 0 | 0 | | <18 | 0 | 0 | | |
| **RHV** | Fontan | -8.34 (14.2) | -36.4 - 19.7 | 0.5579 | 18<= | 2.13 (12.3) | -22.3 - 26.5 | 0.8630 | 0.7018 |
| | Control | 0 | 0 | | <18 | 0 | 0 | | |
| **R_anterior_inferior_hypo** | Fontan | 1.97 (1.7) | -1.38 - 5.31 | 0.2483 | 18<= | 2.21 (1.47) | -0.7 - 5.12 | 0.1353 | 0.6926 |
| | Control | 0 | 0 | | <18 | 0 | 0 | | |
| **R_anterior_superior_hypo** | Fontan | 3.48 (1.55) | 0.43 - 6.54 | 0.0256 | 18<= | 1.86 (1.34) | -0.79 - 4.51 | 0.1684 | 0.3692 |
| | Control | 0 | 0 | | <18 | 0 | 0 | | |
| **R_posterior_hypo** | Fontan | -20.83 (5.51) | -31.7 - -9.95 | 0.0002 | 18<= | -10.31 (4.79) | -19.76 - -0.86 | 0.0326 | 0.0287 |

| | | | | | | | | | |
|---|---|---|---|---|---|---|---|---|---|
| | Control | 0 | 0 | | <18 | 0 | 0 | | |
| **R_tubular_inferior_hypo** | Fontan | 2.52 (5.5) | -8.34 - 13.37 | 0.6476 | 18<= | 3.21 (4.77) | -6.21 - 12.64 | 0.5017 | 0.9369 |
| | Control | 0 | 0 | | <18 | 0 | 0 | | |
| **R_tubular_superior_hypo** | Fontan | 4.52 (4.69) | -4.74 - 13.77 | 0.3365 | 18<= | 5.16 (4.07) | -2.88 - 13.19 | 0.2068 | 0.2802 |
| | Control | 0 | 0 | | <18 | 0 | 0 | | |
| **LHV** | Fontan | -8.48 (12.6) | -33.4 – 16.4 | 0.5020 | 18<= | 9.26 (11.0) | -12.4 - 30.9 | 0.4000 | 0.9815 |
| | Control | 0 | 0 | | <18 | 0 | 0 | | |
| **L_anterior_inferior_hypo** | Fontan | 0.79 (1.36) | -1.89 - 3.48 | 0.5608 | 18<= | 4.07 (1.18) | 1.74 - 6.41 | 0.0007 | 0.9565 |
| | Control | 0 | 0 | | <18 | 0 | 0 | | |
| **L_anterior_superior_hypo** | Fontan | 0.69 (1.29) | -1.85 - 3.23 | 0.5927 | 18<= | 1.86 (1.12) | -0.34 - 4.07 | 0.0976 | 0.6079 |
| | Control | 0 | 0 | | <18 | 0 | 0 | | |
| **L_posterior_hypo** | Fontan | -9.69 (4.89) | -19.35 - -0.03 | 0.0494 | 18<= | -1.71 (4.25) | -10.1 - 6.68 | 0.6882 | 0.6489 |
| | Control | 0 | 0 | | <18 | 0 | 0 | | |
| **L_tubular_inferior_hypo** | Fontan | 2.41 (5.01) | -7.48 - 12.31 | 0.6307 | 18<= | 2.59 (4.35) | -6 - 11.19 | 0.5521 | 0.4815 |
| | Control | 0 | 0 | | <18 | 0 | 0 | | |
| **L_tubular_superior_hypo** | Fontan | -2.69 (4.05) | -10.69 - 5.31 | 0.5075 | 18<= | 2.44 (3.52) | -4.51 - 9.38 | 0.4891 | 0.6668 |
| | Control | 0 | 0 | | <18 | 0 | 0 | | |
| **LHV_norm** | Fontan | 0.26 (0.10) | 0.06 - 0.45 | 0.0125 | 18<= | 0.13 (0.09) | -0.04 - 0.30 | 0.1370 | 0.8563 |
| | Control | 0 | 0 | | <18 | 0 | 0 | | |
| **L_ant_inf_norm** | Fontan | 0.015 | -0.008 - 0.038 | 0.1891 | 18<= | 0.036 (0.01) | 0.016 - 0.056 | 0.0004 | 0.7905 |

| | | | | | | | | | |
|---|---|---|---|---|---|---|---|---|---|
| | | (0.012) | | | | | | | |
| | Control | 0 | 0 | | <18 | 0 | 0 | | |
| **L_ant_sup_norm** | Fontan | 0.025 (0.011) | 0.003 - 0.046 | 0.027 | 18<= | 0.019 (0.01) | -0.0003 - 0.038 | 0.054 | 0.5967 |
| | Control | 0 | 0 | | <18 | 0 | 0 | | |
| **L_post_norm** | Fontan | 0.003 (0.04) | -0.075 - 0.082 | 0.932 | 18<= | 0.003 (0.034) | -0.065 - 0.071 | 0.9363 | 0.5072 |
| | Control | 0 | 0 | | <18 | 0 | 0 | | |
| **L_tub_inf_norm** | Fontan | 0.144 (0.041) | 0.063 - 0.225 | 0.0006 | 18<= | 0.039 (0.036) | -0.032 - 0.11 | 0.2763 | 0.4896 |
| | Control | 0 | 0 | | <18 | 0 | 0 | | |
| **L_tub_sup_norm** | Fontan | 0.068 (0.035) | -0.001 - 0.136 | 0.0541 | 18<= | 0.035 (0.03) | -0.025 - 0.094 | 0.2551 | 0.5054 |
| | Control | 0 | 0 | | <18 | 0 | 0 | | |
| **RHV_norm** | Fontan | 0.24 (0.11) | 0.02 - 0.47 | 0.0316 | 18<= | 0.07 (0.10) | -0.12 - 0.26 | 0.4817 | 0.5460 |
| | Control | 0 | 0 | | <18 | 0 | 0 | | |
| **R_ant_inf_norm** | Fontan | 0.025 (0.014) | -0.003 - 0.053 | 0.0765 | 18<= | 0.021 (0.012) | -0.003 - 0.045 | 0.0876 | 0.7959 |
| | Control | 0 | 0 | | <18 | 0 | 0 | | |
| **R_ant_sup_norm** | Fontan | 0.047 (0.013) | 0.022 - 0.073 | 0.0003 | 18<= | 0.019 (0.011) | -0.003 - 0.041 | 0.0949 | 0.3892 |
| | Control | 0 | 0 | | <18 | 0 | 0 | | |
| **R_post_norm** | Fontan | -0.089 (0.046) | -0.181 - 0.003 | 0.0577 | 18<= | -0.07 (0.04) | -0.15 - 0.009 | 0.0831 | 0.0196 |
| | Control | 0 | 0 | | <18 | 0 | 0 | | |
| **R_tub_inf_norm** | Fontan | 0.132 (0.043) | 0.047 - 0.218 | 0.0027 | 18<= | 0.042 (0.038) | -0.032 - 0.116 | 0.2661 | 0.8526 |
| | Control | 0 | 0 | | <18 | 0 | 0 | | |

| | | | | | | | | | |
|---|---|---|---|---|---|---|---|---|---|
| **R_tub_sup_norm** | Fontan | 0.128 (0.04) | 0.05 - 0.206 | 0.0015 | 18<= | 0.057 (0.034) | -0.011 - 0.125 | 0.0970 | 0.3809 |
| | Control | 0 | 0 | | <18 | 0 | 0 | | |

**Table 4b:** ***Mean Differences in Pituitary and Hypothalamus Volumes Between Groups*** **(WITHOUT interaction term for age)**

| | Group | Estimate (SE) | 95% CI | p | Age_gp | Estimate (SE) | 95% CI | p |
|---|---|---|---|---|---|---|---|---|
| **TPV** | Fontan | 69.4 (20.1) | 29.7 - 109.2 | 0.0007 | 18<= | 47 (21.4) | 4.85 - 89.2 | 0.0291 |
| | Control | 0 | 0 | | <18 | 0 | 0 | |
| **APV** | Fontan | 59.4 (18.7) | 22.4 - 96.5 | 0.0019 | 18<= | 56.2 (19.8) | 17.1 - 95.3 | 0.0051 |
| | Control | 0 | 0 | | <18 | 0 | 0 | |
| **PPV** | Fontan | -4.79 (7.58) | -19.8 - 10.2 | 0.5285 | 18<= | 7.71 (8.00) | -8.10 - 23.5 | 0.3367 |
| | Control | 0 | 0 | | <18 | 0 | 0 | |
| **TPV_norm** | Fontan | 1.17 (0.20) | 0.77 - 1.57 | <.0001 | 18<= | 0.45 (0.21) | 0.03 - 0.88 | 0.0372 |
| | Control | 0 | 0 | | <18 | 0 | 0 | |
| **APV_norm** | Fontan | 0.81 (0.17) | 0.47 - 1.15 | <.0001 | 18<= | 0.61 (0.18) | 0.25 - 0.97 | 0.0011 |
| | Control | 0 | 0 | | <18 | 0 | 0 | |
| **PPV_norm** | Fontan | 0.02 (0.06) | -0.10 - 0.15 | 0.7232 | 18<= | 0.1 (0.07) | -0.04 - 0.23 | 0.1575 |
| | Control | 0 | 0 | | <18 | 0 | 0 | |
| **RHV** | Fontan | -3.91 (8.23) | -20.2 – 12.3 | 0.6357 | 18<= | 5.48 (8.71) | -11.7 – 22.7 | 0.5299 |
| | Control | 0 | 0 | | <18 | 0 | 0 | |
| **R_anterior_inferior_hypo** | Fontan | 1.42 (0.98) | -0.52 - 3.36 | 0.1507 | 18<= | 1.8 (1.04) | -0.25 - 3.85 | 0.0853 |
| | Control | 0 | 0 | | <18 | 0 | 0 | |
| **R_anterior_superior_hypo** | Fontan | 2.35 (0.9) | 0.58 - 4.12 | 0.0097 | 18<= | 1 (0.95) | -0.87 - 2.88 | 0.2923 |
| | Control | 0 | 0 | | <18 | 0 | 0 | |
| **R_posterior_hypo** | Fontan | -10.93 (3.24) | -17.32 - -4.54 | 0.0009 | 18<= | -2.84 (3.42) | -9.6 - 3.91 | 0.4071 |
| | Control | 0 | 0 | | <18 | 0 | 0 | |
| **R_tubular_inferior_hypo** | Fontan | 2.87 (3.18) | -3.41 - 9.16 | 0.3682 | 18<= | 3.48 (3.37) | -3.16 - 10.13 | 0.3024 |
| | Control | 0 | 0 | | <18 | 0 | 0 | |
| **R_tubular_superior_hypo** | Fontan | 0.38 (2.72) | -4.99 - 5.76 | 0.8881 | 18<= | 2.04 (2.88) | -3.64 - 7.73 | 0.4796 |
| | Control | 0 | 0 | | <18 | 0 | 0 | |
| **LHV** | Fontan | -8.72 (7.30) | -23.1 – 5.69 | 0.2339 | 18<= | 9.08 (7.72) | -6.16 – 24.3 | 0.2414 |

| | | | | | | | | |
|---|---|---|---|---|---|---|---|---|
| | Control | 0 | 0 | | <18 | 0 | 0 | |
| L_anterior_inferior_hypo | Fontan | 0.73 (0.79) | -0.82 - 2.29 | 0.3539 | 18<= | 4.03 (0.83) | 2.38 - 5.67 | <.0001 |
| | Control | 0 | 0 | | <18 | 0 | 0 | |
| L_anterior_superior_hypo | Fontan | 0.15 (0.75) | -1.32 - 1.62 | 0.8396 | 18<= | 1.46 (0.79) | -0.1 - 3.01 | 0.0667 |
| | Control | 0 | 0 | | <18 | 0 | 0 | |
| L_posterior_hypo | Fontan | -7.87 (2.84) | -13.47 - -2.27 | 0.0061 | 18<= | -0.34 (3) | -6.26 - 5.58 | 0.9104 |
| | Control | 0 | 0 | | <18 | 0 | 0 | |
| L_tubular_inferior_hypo | Fontan | -0.47 (2.91) | -6.2 - 5.27 | 0.873 | 18<= | 0.42 (3.07) | -5.65 - 6.49 | 0.891 |
| | Control | 0 | 0 | | <18 | 0 | 0 | |
| L_tubular_superior_hypo | Fontan | -1.27 (2.35) | -5.9 - 3.36 | 0.5898 | 18<= | 3.51 (2.48) | -1.39 - 8.41 | 0.1589 |
| | Control | 0 | 0 | | <18 | 0 | 0 | |
| LHV_norm | Fontan | 0.27 (0.06) | 0.15 - 0.39 | <.0001 | 18<= | 0.14 (0.06) | 0.02 - 0.26 | 0.0226 |
| | Control | 0 | 0 | | <18 | 0 | 0 | |
| L_ant_inf_norm | Fontan | 0.018 (0.007) | 0.005 - 0.031 | 0.0088 | 18<= | 0.038 (0.007) | 0.024 - 0.052 | <.0001 |
| | Control | 0 | 0 | | <18 | 0 | 0 | |
| L_ant_sup_norm | Fontan | 0.02 (0.006) | 0.007 - 0.033 | 0.0022 | 18<= | 0.015 (0.007) | 0.002 - 0.028 | 0.0278 |
| | Control | 0 | 0 | | <18 | 0 | 0 | |
| L_post_norm | Fontan | 0.025 (0.023) | -0.021 - 0.07 | 0.2813 | 18<= | 0.019 (0.024) | -0.029 - 0.067 | 0.437 |
| | Control | 0 | 0 | | <18 | 0 | 0 | |
| L_tub_inf_norm | Fontan | 0.121 (0.024) | 0.074 - 0.168 | <.0001 | 18<= | 0.022 (0.025) | -0.028 - 0.071 | 0.3946 |
| | Control | 0 | 0 | | <18 | 0 | 0 | |
| L_tub_sup_norm | Fontan | 0.087 (0.02) | 0.047 - 0.126 | <.0001 | 18<= | 0.049 (0.021) | 0.007 - 0.091 | 0.0235 |
| | Control | 0 | 0 | | <18 | 0 | 0 | |
| RHV_norm | Fontan | 0.30 (0.07) | 0.17 - 0.43 | <.0001 | 18<= | 0.11 (0.07) | -0.03 - 0.25 | 0.1103 |
| | Control | 0 | 0 | | <18 | 0 | 0 | |
| R_ant_inf_norm | Fontan | 0.022 (0.008) | 0.006 - 0.038 | 0.0073 | 18<= | 0.019 (0.009) | 0.002 - 0.036 | 0.0309 |

| | | | | | | | | |
|---|---|---|---|---|---|---|---|---|
| | Control | 0 | 0 | | <18 | 0 | 0 | |
| **R_ant_sup_norm** | Fontan | 0.038 (0.007) | 0.024 - 0.053 | <.0001 | 18<= | 0.012 (0.008) | -0.004 - 0.027 | 0.1321 |
| | Control | 0 | 0 | | <18 | 0 | 0 | |
| **R_post_norm** | Fontan | 0.0003 (0.027) | -0.054 - 0.054 | 0.9909 | 18<= | -0.003 (0.029) | -0.06 - 0.054 | 0.9139 |
| | Control | 0 | 0 | | <18 | 0 | 0 | |
| **R_tub_inf_norm** | Fontan | 0.139 (0.025) | 0.089 - 0.188 | <.0001 | 18<= | 0.047 (0.027) | -0.005 - 0.099 | 0.0787 |
| | Control | 0 | 0 | | <18 | 0 | 0 | |
| **R_tub_sup_norm** | Fontan | 0.099 (0.023) | 0.054 - 0.145 | <.0001 | 18<= | 0.036 (0.024) | -0.012 - 0.084 | 0.1400 |
| | Control | 0 | 0 | | <18 | 0 | 0 | |

***Table 4c: Mean Differences in Pituitary and Hypothalamus Volumes Between Groups with Multivariable Regression for Age (continuous)***

| | | | Estimate (SE) | 95% CI | p |
|---|---|---|---|---|---|
| **TPV_norm** | Group | Fontan | 4.79 (0.45) | 3.90 - 5.68 | <.0001 |
| | Group | Control | 3.79 (0.44) | 2.92 - 4.66 | |
| | Age_yr | | 0.025 (0.018) | -0.010 - 0.061 | **0.0294** |
| | Age_yr*Group | Fontan | 0.006 (0.026) | -0.045 - 0.057 | 0.8165 |
| | Age_yr*Group | Control | 0 | | |
| **RHV_norm** | Group | Fontan | 3.43 (0.14) | 3.14 - 3.71 | <.0001 |
| | Group | Control | 3.16 (0.14) | 2.89 - 3.44 | |
| | Age_yr | | 0.002 (0.006) | -0.009 - 0.014 | 0.4581 |
| | Age_yr*Group | Fontan | 0.002 (0.008) | -0.015 - 0.018 | 0.8505 |
| | Age_yr*Group | Control | 0 | | |
| **LHV_norm** | Group | Fontan | 3.47 (0.13) | 3.22 - 3.73 | <.0001 |
| | Group | Control | 3.18 (0.13) | 2.93 - 3.42 | |
| | Age_yr | | 0.006 (0.005) | -0.004 - 0.016 | 0.1220 |
| | Age_yr*Group | Fontan | -0.001 (0.007) | -0.016 - 0.014 | 0.8832 |
| | Age_yr*Group | Control | 0 | | |

**Table 5:** ***Associations between clinical and demographic variables and pituitary and hypothalamus volumes***

| | | Estimate (SE) | 95% CI | p |
|---|---|---|---|---|
| **TPV_norm** | Time_Fontan_MRI_yr | 0.0521 (0.0252) | 0.0020 - 0.1022 | 0.0415 |
| | Age1st_surgery_days | -0.0005 (0.0004) | -0.0014 - 0.0004 | 0.3056 |
| | BMI | 0.0307 (0.0338) | -0.0365 - 0.0978 | 0.3661 |
| | Ht_centile | 0.0069 (0.0056) | -0.0043 - 0.018 | 0.2242 |
| **APV_norm** | Time_Fontan_MRI_yr | 0.0677 (0.0206) | 0.0266 - 0.1088 | 0.0016 |
| | Age1st_surgery_days | -0.0001 (0.0004) | -0.001 - 0.0007 | 0.7562 |
| | BMI | 0.0483 (0.0318) | -0.0153 - 0.1118 | 0.1338 |
| | Ht_centile | 0.0127 (0.0051) | 0.0024 - 0.023 | 0.0163 |
| **PPV_norm** | Time_Fontan_MRI_yr | 0.0141 (0.0075) | -0.001 - 0.0292 | 0.0668 |
| | Age1st_surgery_days | 0.0001 (0.0001) | -0.0002 - 0.0004 | 0.4829 |
| | BMI | 0.0120 (0.0112) | -0.0103 - 0.0343 | 0.2846 |
| | Ht_centile | 0.0028 (0.0018) | -0.0009 - 0.0065 | 0.1370 |
| **LHV_norm** | Time_Fontan_MRI_yr | 0.0154 (0.0061) | 0.0034 - 0.0275 | 0.0129 |
| | Age1st_surgery_days | 0 (0.0001) | -0.0003 - 0.0002 | 0.7041 |
| | BMI | 0.0054 (0.0083) | -0.0111 - 0.0218 | 0.5161 |
| | Ht_centile | 0.0016 (0.0014) | -0.0011 - 0.0043 | 0.2478 |
| **RHV_norm** | Time_Fontan_MRI_yr | 0.0127 (0.0059) | 0.0009 - 0.0244 | 0.0354 |
| | Age1st_surgery_days | -0.0001 (0.0001) | -0.0003 - 0.0002 | 0.6068 |
| | BMI | -0.0035 (0.0080) | -0.0195 - 0.0124 | 0.6594 |
| | Ht_centile | 0.0007 (0.0013) | -0.0019 - 0.0034 | 0.5815 |

**Figure 5: Normalized Pituitary Volumes with Duration of Fontan Circulation** (ie. Time between Fontan completion and MRI)

*Note that normalized pituitary volumes are a percentage of total brain volume scaled by a factor of 100.*

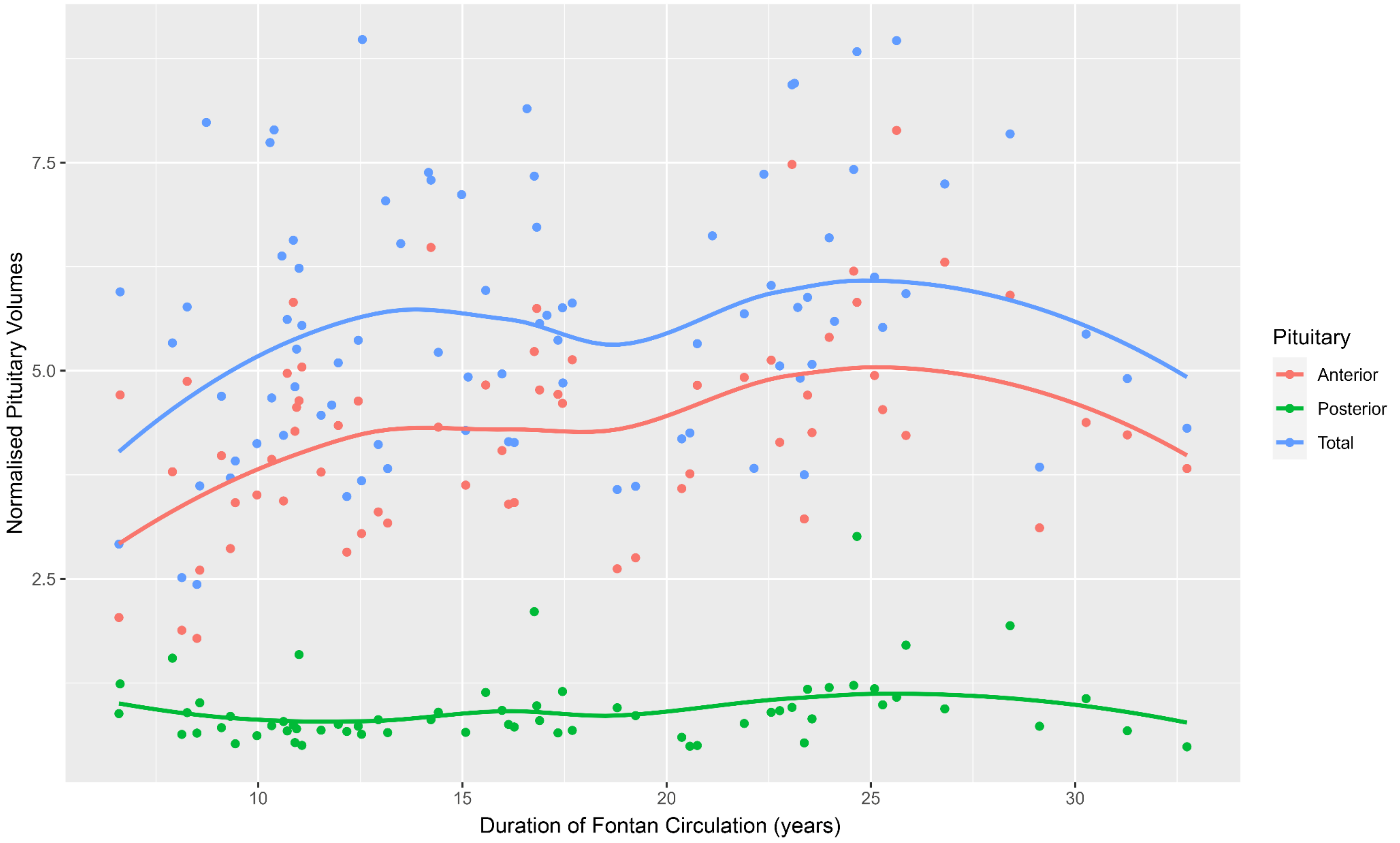

**Table 6:** ***Associations between sex and pituitary and hypothalamus volumes (male vs female)***

| | | Fontan | | | Control | | | | | |
|---|---|---|---|---|---|---|---|---|---|---|
| **Variable** | Sex | N | Mean (SD) | 95% CI | N | Mean (SD) | 95% CI | p-value for Group | p-value for Sex | p-value for Group*Sex |
| **TPV_norm** | F | 37 | 6.46 (1.32) | 6.02 - 6.9 | 37 | 5.14 (0.9) | 4.84 - 5.44 | <.0001 | <.0001 | 0.4599 |
| | M | 48 | 4.86 (1.32) | 4.47 - 5.24 | 49 | 3.8 (0.89) | 3.54 - 4.05 | | | |
| **APV_norm** | F | 24 | 5.14 (1.08) | 4.69 - 5.6 | 37 | 4.18 (0.77) | 3.92 - 4.44 | <.0001 | <.0001 | 0.6868 |
| | M | 40 | 3.81 (1.02) | 3.48 - 4.13 | 49 | 2.96 (0.75) | 2.75 - 3.18 | | | |
| **PPV_norm** | F | 24 | 1 (0.56) | 0.77 - 1.24 | 37 | 0.96 (0.43) | 0.82 - 1.11 | 0.6946 | 0.0458 | 0.9363 |
| | M | 40 | 0.87 (0.32) | 0.76 - 0.97 | 49 | 0.84 (0.31) | 0.75 - 0.93 | | | |
| **LHV_norm** | F | 37 | 3.66 (0.39) | 3.53 - 3.79 | 37 | 3.37 (0.37) | 3.24 - 3.49 | <.0001 | 0.0789 | 0.7497 |
| | M | 49 | 3.54 (0.36) | 3.43 - 3.64 | 49 | 3.28 (0.42) | 3.16 - 3.4 | | | |
| **RHV_norm** | F | 37 | 3.56 (0.45) | 3.41 - 3.71 | 37 | 3.33 (0.41) | 3.2 - 3.47 | <.0001 | 0.0287 | 0.3241 |
| | M | 49 | 3.48 (0.29) | 3.40 - 3.56 | 49 | 3.12 (0.52) | 2.97 - 3.27 | | | |

**Table 7:** ***Between group comparisons of the number of cardiac surgeries prior to Fontan completion, and pituitary and hypothalamus volumes*****.**

| Variable | No_operations_prior_Fontan | N | Mean (SD) | p |
|---|---|---|---|---|
| **TPV** | 0 | 1 | 674.0 | 0.9536 |
| | 1 | 25 | 604.4 (129.5) | |
| | 2 | 32 | 594.2 (147.8) | |
| | 3 | 16 | 580.1 (125.7) | |
| | 4 | 11 | 603.9 (134.7) | |
| **APV** | 0 | 1 | 544.0 | 0.6638 |
| | 1 | 18 | 496.3 (137.5) | |
| | 2 | 27 | 467.5 (117.1) | |
| | 3 | 11 | 452.0 (110.5) | |
| | 4 | 7 | 522.4 (85.7) | |
| **PPV** | 0 | 1 | 130.0 | 0.5140 |
| | 1 | 18 | 100.9 (38.1) | |
| | 2 | 27 | 105.4 (55.4) | |
| | 3 | 11 | 85.5 (20.7) | |
| | 4 | 7 | 123.1 (57.2) | |
| **LHV** | 0 | 1 | 363.0 | 0.8086 |
| | 1 | 25 | 393.8 (42.5) | |
| | 2 | 33 | 396.4 (49.7) | |
| | 3 | 16 | 389.4 (49.9) | |
| | 4 | 11 | 381.3 (37.4) | |
| **RHV** | 0 | 1 | 371.0 | 0.7769 |
| | 1 | 25 | 386.2 (45.8) | |
| | 2 | 33 | 386.1 (48.1) | |
| | 3 | 16 | 373.1 (52.7) | |

| | | | |
|---|---|---|---|
| | 4 | 11 | 390.5 (51.1) |

**Table 8:** ***Between group comparisons of predominant ventricular morphology and pituitary and hypothalamus volumes***

| Variable | Predominant_ventricular_morphology | N | Mean (SD) | p |
|---|---|---|---|---|
| **TPV** | Biventricular | 6 | 610.5 (90.3) | 0.8467 |
| | Indeterminate | 5 | 517.6 (143.3) | |
| | Left | 49 | 601.3 (140.0) | |
| | Right | 25 | 600.2 (131.2) | |
| **APV** | Biventricular | 5 | 512.6 (96.2) | 0.9952 |
| | Indeterminate | 5 | 422.0 (116.9) | |
| | Left | 37 | 482.0 (117.5) | |
| | Right | 17 | 483.6 (129.1) | |
| **PPV** | Biventricular | 5 | 84.0 (6.4) | 0.6936 |
| | Indeterminate | 5 | 96.0 (31.6) | |
| | Left | 37 | 105.6 (51.8) | |
| | Right | 17 | 104.9 (44.9) | |
| **LHV** | Biventricular | 7 | 396.9 (38.5) | 0.5372 |
| | Indeterminate | 5 | 409.0 (37.1) | |
| | Left | 49 | 396.5 (45.0) | |
| | Right | 25 | 378.5 (49.1) | |
| **RHV** | Biventricular | 7 | 394.3 (32.8) | 0.8891 |
| | Indeterminate | 5 | 406.6 (33.6) | |
| | Left | 49 | 385.0 (52.2) | |
| | Right | 25 | 375.0 (44.5) | |

**Table 9:** ***Between group comparisons of Fontan Type and normalized pituitary and hypothalamus volumes***

| | Fontan_Type | N | Mean (SD) | p |
|---|---|---|---|---|
| **TPV** | AP | 9 | 653.2 (133.7) | 0.7656 |
| | AP converted to ECC | 1 | 454.0 | |
| | ECC | 56 | 584.5 (130.9) | |
| | LT | 17 | 619.1 (146) | |
| | LT converted to ECC | 1 | 656.0 | |
| **APV** | AP | 9 | 541.7 (111.9) | 0.9217 |
| | AP converted to ECC | 1 | 403.0 | |
| | ECC | 42 | 460 (115.9) | |
| | LT | 10 | 521.9 (125.6) | |
| | LT converted to ECC | 1 | 525.0 | |
| **PPV** | AP | 9 | 111.6 (39) | 0.7189 |
| | AP converted to ECC | 1 | 51.0 | |
| | ECC | 42 | 99.2 (36.6) | |
| | LT | 10 | 114.8 (83.2) | |
| | LT converted to ECC | 1 | 131.0 | |
| **LHV** | AP | 9 | 393.8 (45.2) | 0.9896 |
| | AP converted to ECC | 1 | 339.0 | |
| | ECC | 57 | 391.6 (48.4) | |
| | LT | 17 | 391.1 (38.1) | |
| | LT converted to ECC | 1 | 433.0 | |
| **RHV** | AP | 9 | 386.9 (47.5) | 0.9695 |
| | AP converted to ECC | 1 | 319.0 | |
| | ECC | 57 | 382.7 (49.8) | |
| | LT | 17 | 384 (40.6) | |
| | LT converted to ECC | 1 | 445.0 | |
